# Assessing the Youth Safety Lab Program Through a Health Equity Lens

**DOI:** 10.1101/2025.09.09.25335127

**Authors:** Qizhi Mao, Shaelyn Fitzpatrick, Brandy Tanenbaum

## Abstract

**Background:** Injury is the leading cause of death among youth in Canada. This study aimed to evaluate the effectiveness of the Youth Safety Lab (YSL) program in improving injury prevention knowledge and skills among high school students in the Greater Toronto Area, with attention to equity across sociodemographic groups.

**Methods:** A retrospective longitudinal study involved 1,805 students from over 50 high schools at three time points: before the program, immediately after, and three months later. The survey, based on the Health Action Process Approach (HAPA), assessed five aspects related to injury prevention. Wilcoxon signed-rank tests and regression analyses were used to examine changes and disparities in outcomes.

**Results:** Scores across all five sections showed significant improvement following the intervention, with the greatest gains observed in bleeding injury response. Black students were underrepresented in follow-up participation, and female students exhibited greater improvements than male students in bleeding injury response, indicating potential disparities in engagement and outcomes.

**Conclusions:** The YSL program effectively enhances injury prevention awareness among high school students in the GTA. However, disparities in participation and outcomes highlight the need for more inclusive and responsive injury prevention programs.

**What is already known on this topic:**

- The P.A.R.T.Y. (Prevent Alcohol and Risk-Related Trauma in Youth) program has been demonstrated to effectively improve students’ risk awareness.
- Injury risk and its impact vary unequally among different population groups.

**What this study adds:**

- The YSL program demonstrated notable improvements in injury prevention knowledge and skills, especially in bleeding injury response.
- Disparities in access to and outcomes of health education interventions persist among diverse demographic groups.

**How might this study affect research, practice, or policy:**

- Reinforces the value of injury prevention programs like the YSL and emphasizes the need to explore sustainable models for access, funding, and scalability to better reach priority populations. Highlights the need to design more inclusive and responsive injury prevention programs that address barriers to participation and ensure equitable benefits for diverse youth populations.

## 1. Introduction

Injury-related mortality constitutes a substantial burden on global public health. According to the World Health Organization, injuries account for over 4.4 million deaths annually worldwide, comprising nearly 8% of all deaths^1^. In Canada, injuries are the leading cause of death among individuals aged 1 to 44, with 74% of these deaths resulting from unintentional and intentional injuries^2^. In 2018, the total economic cost of injuries for children (aged 14 and under) and youth (aged 15 to 24) was $2.9 billion CAD and $4.6 billion CAD, respectively^3^.

Youth experience neurological changes during puberty that affect risk perception, leading to poor decision-making, diminished impulse control, and difficulties with emotional regulation^4,5^. Additionally, there are significant inequities in the risk and types of injuries experienced by youth^6^. These disparities are shaped by intersecting factors such as, income, race, gender, and geographic location. For example, youth from lower socioeconomic status (SES) are more likely to experience alcohol abuse, violence, and bleeding injuries from fights, while those with higher SES are more likely to experience sports-related injuries^7,8^. Identifying populations that are underserved by injury prevention programs is crucial to ensure that interventions are inclusive, equitable, and effective across diverse groups.

School and community-based health education plays a key role in reducing high-risk injury behaviors among youth^4^. The Youth Safety Lab (YSL) is an injury prevention initiative that incorporates both primary and secondary prevention education through participation in the P.A.R.T.Y. Program (Prevent Alcohol and Risk Related Trauma in Youth) and STOP THE BLEED® training. The P.A.R.T.Y. Program (www.partyprogram.com) was established in 1986 at Sunnybrook Health Sciences Centre, the first and largest trauma center in Canada, located in Toronto, Ontario. Aimed at youth aged 15 and older, the program provides a one-day health education experience designed to increase awareness and knowledge about traumatic injuries. By addressing topics such as impairment, motor vehicle collisions, and other risk-related behaviors, the program serves as an effective form of primary prevention to reduce the occurrence of traumatic injuries. STOP THE BLEED® (www.stopthebleed.org) is a national awareness campaign and training program that was launched in 2015 by the American College of Surgeons. Developed in response to the 2012 Sandy Hook Elementary School shooting, STOP THE BLEED® training empowers bystanders to act quickly and effectively in bleeding emergencies. Since 2017, Sunnybrook Health Sciences Center has been delivering this training to community members across the GTA (Greater Toronto Area). Through hands-on sessions, participants learn life-saving techniques, such as applying pressure, packing deep wounds, and using tourniquets.

This study aims to evaluate the effectiveness of the YSL program in enhancing injury prevention awareness among high school students in the GTA, with a particular focus on bleeding injuries. Additionally, it investigates inequities in program participation and the benefits gained across groups.

## 2. Method

### 2.1 Design and Participant Recruitment

This retrospective longitudinal study was conducted at Sunnybrook Health Sciences Centre. The participants were students from over 50 high schools in the GTA that voluntary took part in Sunnybrook’s Youth Safety Lab. Accompanied by teachers or group leaders, students attended a one-day injury awareness and prevention education program.

Before the program began, students were invited to complete a pre-survey by scanning a QR code, which also collected relevant sociodemographic information. Survey responses were collected through REDCap data collection software. The health education sessions were delivered by specialists from the Centre for Injury Prevention and lasted approximately six hours.

Participants received the second questionnaire (post-survey) on the same day of the workshop and the third questionnaire (post-postsurvey) after three months. The three surveys were identical in content, while sociodemographic information collected only in the pre-survey. The latter two surveys were sent via email, and students were able to complete and submit them online.

### 2.2 Measures

Participants provided sociodemographic information including whether they were born in Canada (and if not, how long since they arrived in Canada), whether they self-identify as Indigenous (including First Nations, Métis, and Inuk/Inuit), ethnic or cultural background, whether they self-identify as a person with a disability, sex assigned at birth, gender identity, whether they self-identify as transgender, and sexual or asexual orientation. Participants could select multiple options for questions related to ethnic or cultural background and gender identity. To protect participants’ rights, all questions included response options such as “Do not know” and “Prefer not to answer,” which were treated as “Not provided” during analysis.

The questionnaire was designed based on the Health Action Process Approach (HAPA) framework and comprised five sections^9,10^. The first section, Injury Awareness, examines participants’ risk perception and self-efficacy related to injury, consisting of 5 items. The second and third sections, Scenario-Based Prevention Skills and Peer-Related Prevention Response, focus on the volitional phase of behavior change, particularly action planning. These sections assess participants’ confidence and ability to develop coping strategies in various injury-related scenarios, both when alone and when interacting with peers, and together include 9 items. The fourth section, Bleeding Injury Response, evaluates participants’ knowledge, skills, and confidence in responding to bleeding injury situations, with a total of 3 items. The fifth section, Injury Concern Levels, investigates participants’ level of concern regarding different types and locations of injuries, and contains 6 items. All items are measured using a 5-point Likert scale, with a maximum total score of 115 points. Scores are calculated individually for each item and section. Higher scores indicate more positive outcomes, such as greater confidence, knowledge, and awareness related to injury prevention and response.

### 2.3 Statistical analysis

Statistical analysis and data visualization were performed using SAS 9.4 and R version 4.4.1, with the significance level set at 0.05 for all tests. Descriptive analyses were conducted to characterize the participants’ sociodemographic information. The Wilcoxon Signed-Rank Test was used to analyze the changes in scores between pre-test and post-test, as well as between pre-test and the post-post survey. Simple linear regression models were constructed, and variables with p < 0.1 were included in the multiple linear regression model to explore the effects of participants’ sociodemographic characteristics on the section 4 of the survey: Bleeding Injury Response.

## 3. Result

### 3.1 Participant Characteristics

Between December 2022 and April 2025, a total of 2,663 high school students attended the YSL. Of these, 1,805 (68%) consented to and participated in the pre-survey, which included the collection of sociodemographic information. Among them, 650 students (36%) participated in the post-survey after the program, and 212 students (12%) completed the post post-survey after three months. Overall, excluding those who did not report their racial or cultural background, the proportion of black students were significantly lower in the post-survey compared to that in the pre-survey (12.3% vs. 16.1%; p <.001), while no significant differences were found in other sociodemographic characteristics (see Table 1).

**Table 1.** Sociodemographic Characteristics of Survey Respondents.

| Description |  | Pre |  | Post |  | Chi-square | P-Value |
| --- | --- | --- | --- | --- | --- | --- | --- |
|  |  | N | % | N | % |  |  |
| Time to arrive | **Born in Canada** | 1211 | 67.1 | 450 | 69.2 | 1.96 | 0.74 |
|  | **Less than 5 years** | 175 | 9.7 | 59 | 9.1 |  |  |
|  | **5-9 years** | 191 | 10.6 | 64 | 9.8 |  |  |
|  | **More than 10 years** | 194 | 10.7 | 69 | 10.6 |  |  |
|  | **Not provided** | 34 | 1.9 | 8 | 1.2 |  |  |
| Do you identify as Indigenous? | **Yes** | 36 | 2.0 | 12 | 1.8 | 2.4 | 0.30 |
|  | **No** | 1695 | 93.9 | 620 | 95.4 |  |  |
|  | **Not provided** | 74 | 4.1 | 18 | 2.8 |  |  |
| What is your ethnic or cultural background? | **Middle Eastern, Arab or West Asian** | 208 | 11.5 | 67 | 10.3 | 407.0 | *** |
|  | **Black** | 290 | 16.1 | 80 | 12.3 |  |  |
|  | **East Asian** | 192 | 10.6 | 87 | 13.4 |  |  |
|  | **Jewish** | 51 | 2.8 | 22 | 3.4 |  |  |
|  | **Latin American** | 94 | 5.2 | 29 | 4.5 |  |  |
|  | **South Asian** | 296 | 16.4 | 102 | 15.7 |  |  |
|  | **Southeast Asian** | 324 | 18.0 | 105 | 16.2 |  |  |
|  | **White** | 453 | 25.1 | 196 | 30.2 |  |  |
|  | **Another Race** | 29 | 1.6 | 14 | 2.2 |  |  |
|  | **Prefer not to answer** | 54 | 3.0 | 22 | 3.4 |  |  |
|  | **Do not know** | 25 | 1.4 | 7 | 1.1 |  |  |
| Do you identify as a person with disability? | **Yes** | 54 | 3.0 | 11 | 1.7 | 3.2 | 0.20 |
|  | **No** | 1636 | 90.6 | 595 | 91.5 |  |  |
|  | **Not provided** | 115 | 6.4 | 44 | 6.8 |  |  |
| What is your gender identity? | **Man** | 487 | 27.0 | 172 | 26.5 | 1.09 | 0.58 |
|  | **Woman** | 1231 | 68.2 | 440 | 67.7 |  |  |
|  | **Others or not provided** | 117 | 6.5 | 50 | 7.7 |  |  |
| Sex at Birth | **Male** | 495 | 27.4 | 176 | 27.1 | 1.14 | 0.57 |
|  | **Female** | 1287 | 71.3 | 469 | 72.2 |  |  |
|  | **Not provided** | 23 | 1.3 | 5 | 0.8 |  |  |
| Transgender | **Yes** | 31 | 1.7 | 14 | 2.2 | 0.7 | 0.69 |
|  | **No** | 1773 | 98.2 | 620 | 95.4 |  |  |
|  | **Not provided** | 41 | 2.3 | 16 | 2.5 |  |  |
| Which category(ies) best describe your sexual Asexual orientation? | **Straight/Heterosexual** | 1322 | 73.2 | 464 | 71.4 | 0.8 | 0.66 |
|  | **Others** | 228 | 12.6 | 87 | 13.4 |  |  |
|  | **Not provided** | 255 | 14.1 | 99 | 15.2 |  |  |

\*\*\* Significant differences between pre- and post-Survey ( $p \leq 0.05$ ).

### 3.2 Change in injury-related knowledge and health behaviors

Descriptive statistics for the pre- and post-surveys are shown in Table 2. All sections of the questionnaire showed significant improvements from before (pre-survey) to after (post-survey) the health education program (p <.001). As shown in Table 2, injury prevention and awareness knowledge improved across all five dimensions. At the item level, significant improvements were observed in all 23 questions related to injury prevention and awareness (see Figure 1).

**Table 2.**
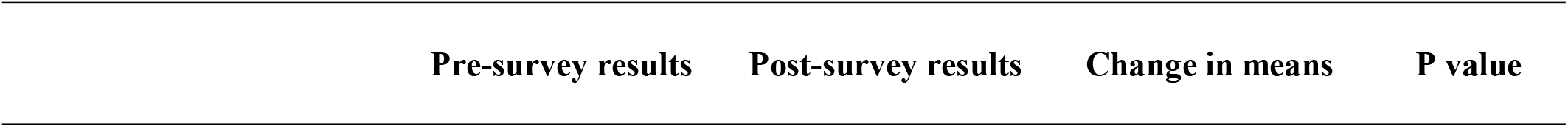
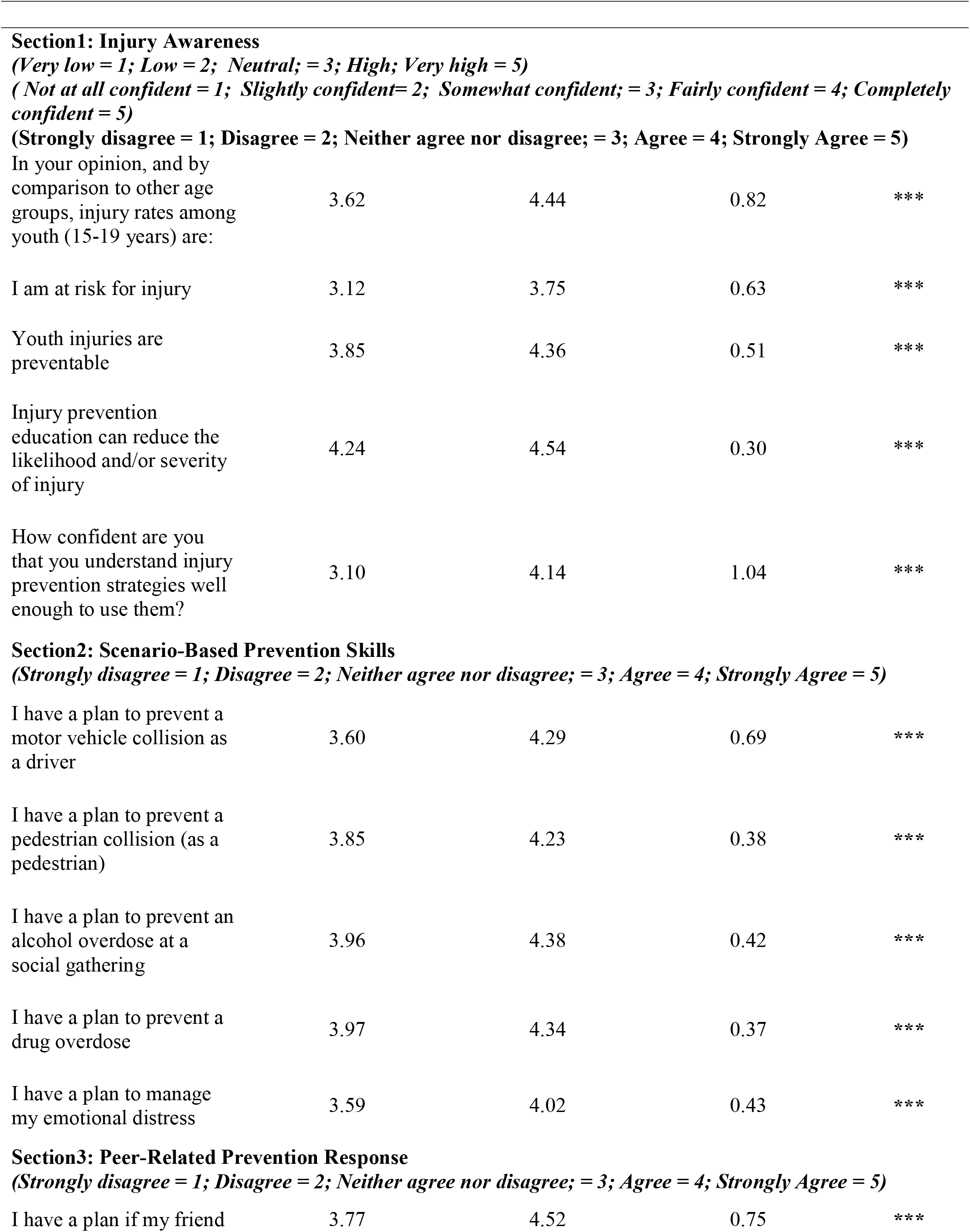
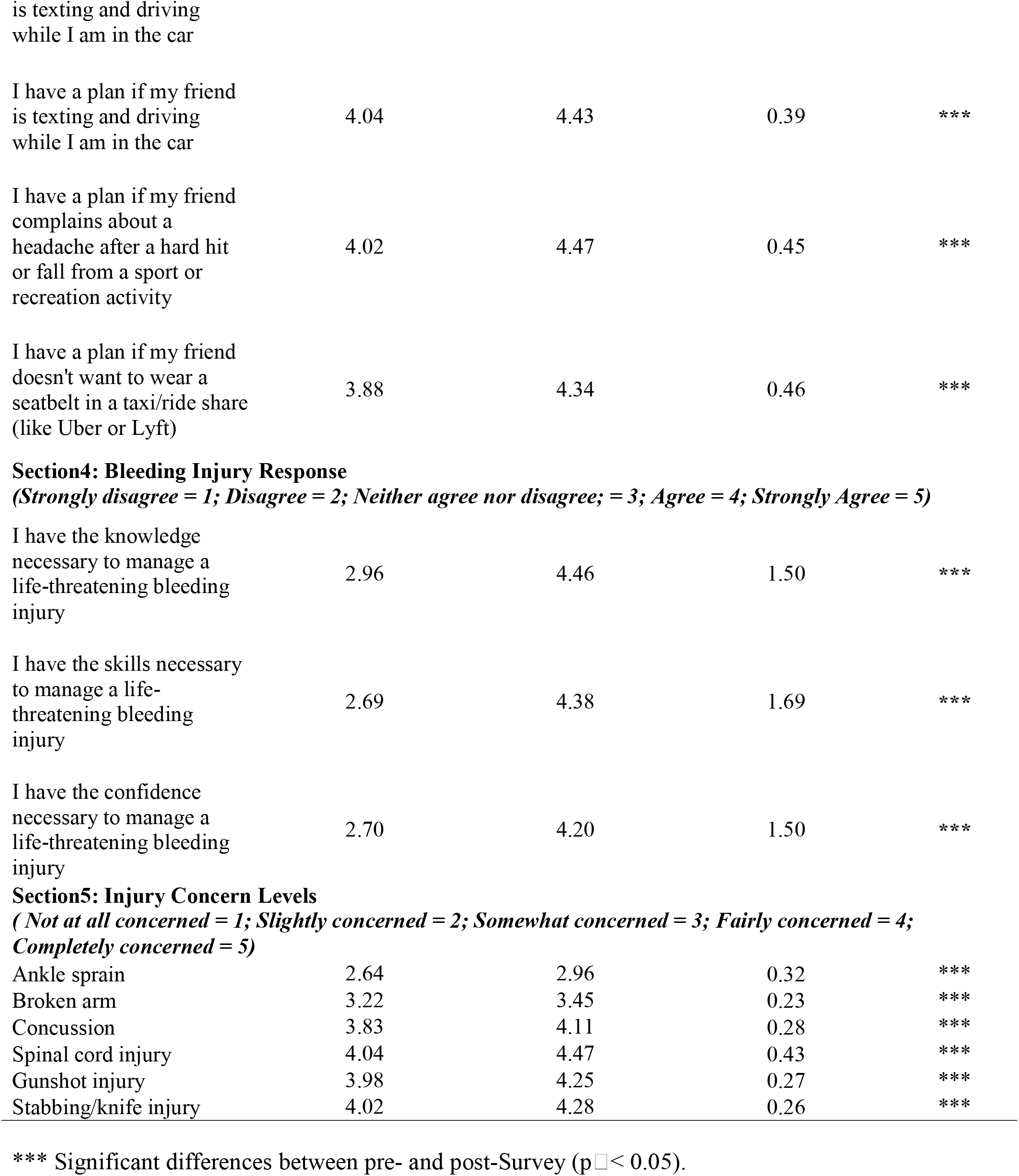
Changes in scores across five sections in the overall sample between the Pre- and Post-Survey.

**Figure 1.**
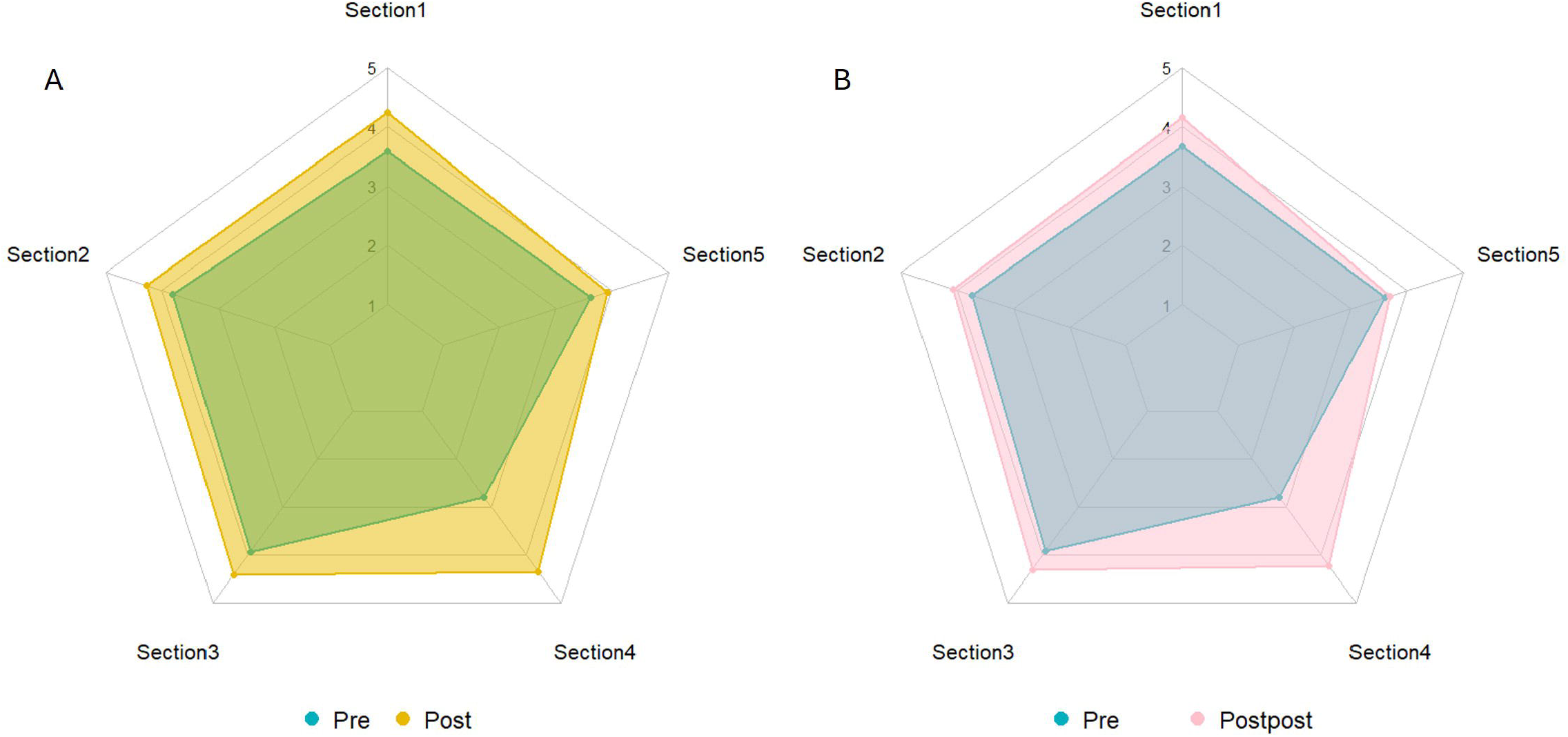
A. Comparison between Pre- and Post-Survey. B. Comparison between Pre- and Post post-Survey.

The most distinct difference between the pre- and post-surveys was observed in Section 4: Bleeding Injury Response. All three questions in this section showed a score increase of more than one point (IQR = 4).

In Section 1: Injury Awareness, although each question showed a significant difference between the pre- and post-surveys, only the item “Confidence level in injury prevention strategy use” had an average score increase of more than 1 point. In the sections Scenario-Based Prevention Skills and Peer-Related Prevention Response, which include 9 questions, none of them showed a difference greater than 1 point, and two of the questions had average scores above 4 in the pre-survey. In the Injury Concern Levels section, the overall score improvement was relatively small, with 4 out of 6 questions showing an average increase of less than 0.3 points. Overall, the post-post survey showed similar results (see Figure 1).

### 3.3 Linear regression analysis

In terms of the disparities present in Section 4: Bleeding Injury Response between the pre- and post-surveys, simple linear regression analysis showed that arriving in Canada 5–9 years ago (B = 0.85; p = 0.03), White ethnicity (B = −0.73; p = 0.04), and female (B = 0.75; p = 0.00) had significant effects on changes in injury prevention knowledge and awareness. Multiple linear regression analysis identified female (B = 0.75; p = 0.006) as a significant and independent factor (see Table 3).

**Table 3.**
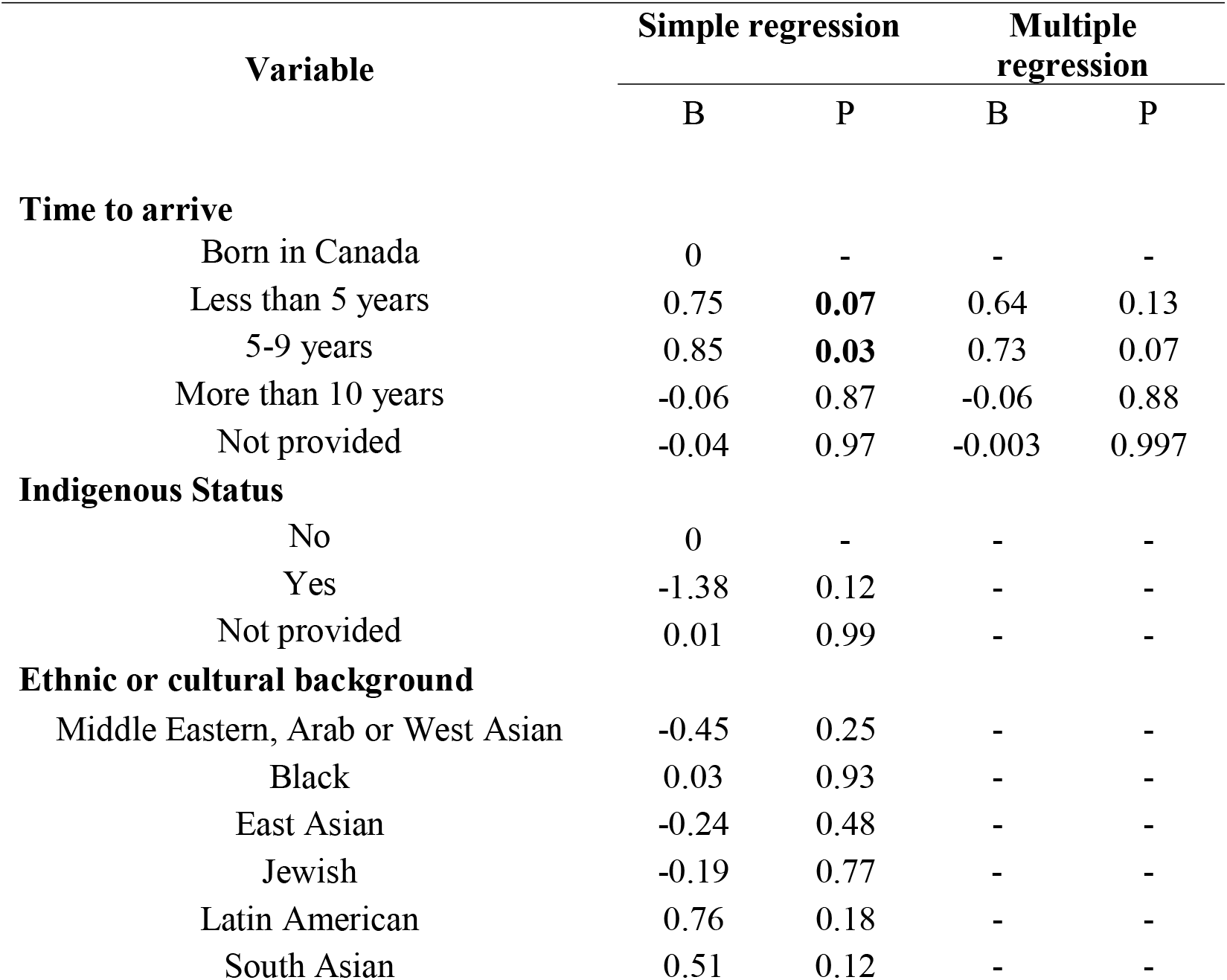
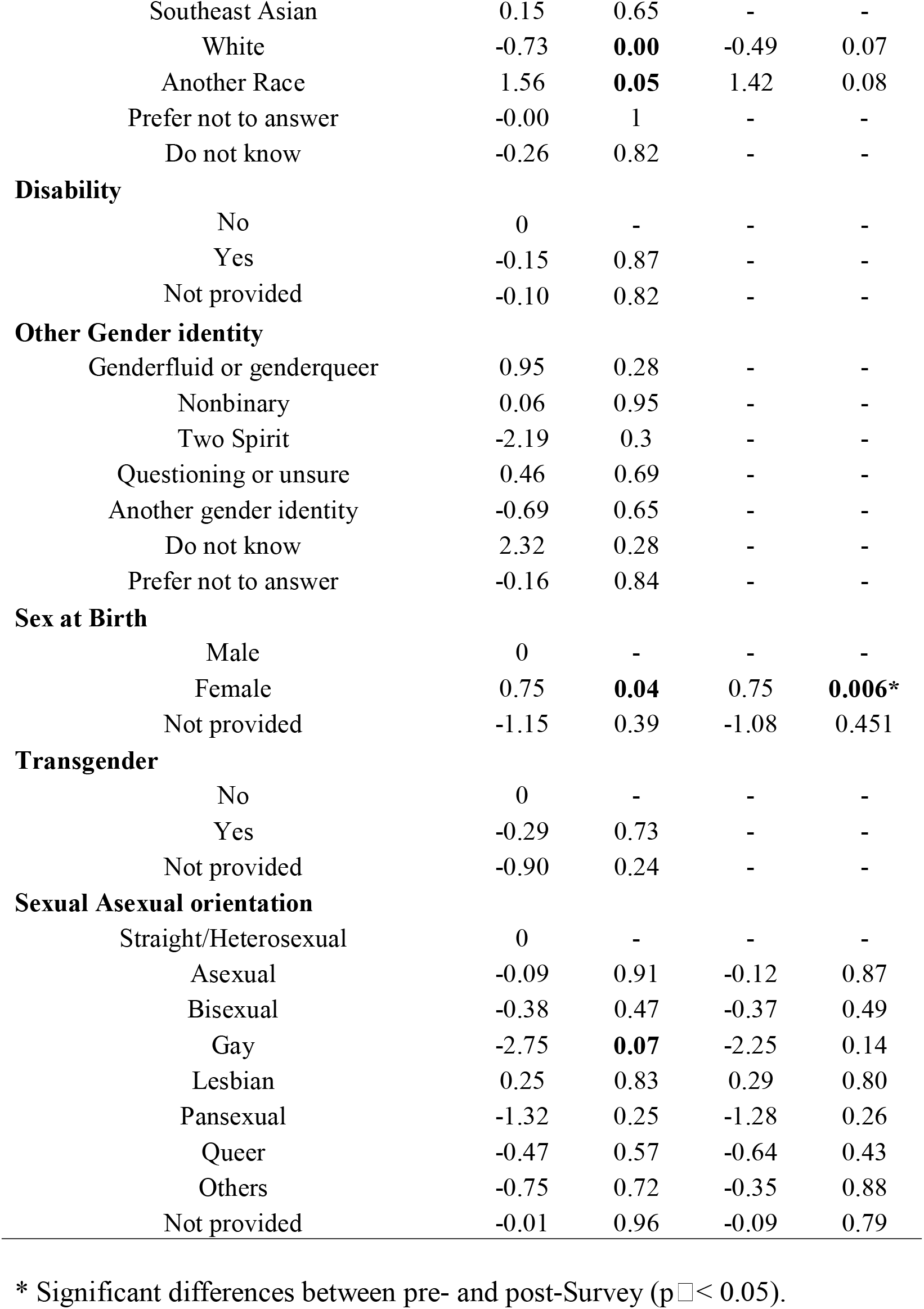
Simple and multiple regression analyses of Section 4 post-survey scores in the overall sample.

## 4. Discussion and Conclusions

The P.A.R.T.Y. program has been shown to be effective in enhancing students’ risk awareness, particularly in relation to alcohol, drug use, and various risk-related behaviors^11,12^. A Germany study indicated that participation in the program helped students develop an understanding of the dangers associated with road traffic and appropriate safety behaviors^13^. However, previous studies have largely overlooked bleeding injuries and the unequal distribution of benefits among different groups.

This study evaluated both short- and long-term effects of Sunnybrook’s YSL program, across more than fifty high schools in Toronto. All five sections of the questionnaire showed improvements after the intervention, indicating that this one-day health education program effectively enhanced students’ risk perception, coping strategies, and intentions to adopt safer behaviors. Notably, Section 4 demonstrated an average increase of more than one point, suggesting a substantial improvement in students’ knowledge and confidence in managing bleeding injuries. The relatively low baseline scores observed in this section reflected the lack of emphasis on bleeding injury in other educational programs. The YSL program successfully addressed this gap by incorporating STOP THE BLEED® training.

In comparison, the first three sections, although statistically significant, showed more modest improvements, with only one item “Confidence in using injury prevention strategies” showing an average increase of over one point. This limited change may be attributable to relatively high baseline scores across several items, suggesting that students already had a foundational awareness of general injury risks. It also highlights a potential need for injury prevention specialists to reconsider the framing of assessment items to more objectively evaluate educational impact. Section 5, Injury Concern Levels, showed the smallest change, which may reflect the program’s intentional focus on more severe, life-altering injuries rather than common minor injuries (e.g., ankle sprains). This suggests that students appropriately prioritized traumatic injury concerns, aligning with the program’s objectives.

From a health equity lens, the study identified a lower proportion of Black students participating in the post-intervention survey compared to other racial and cultural groups. This disparity may indicate that the intervention is not resonating with or reaching Black students as effectively as it is with other groups, whether due to cultural relevance, perceived safety, or lack of trust in health-related programming. Additionally, it may indicate that Black students encounter additional barriers to accessing follow-up health education opportunities, potentially stemming from constraints such as limited discretionary time, inadequate internet access, or broader socioeconomic challenges. Linear regression analysis further revealed that male students showed significantly less improvement in Bleeding Injury Response compared to female students. Moreover, over 70% of the students who voluntarily participated in the program were female, highlighting a significant gender disparity in engagement, which has been consistently observed in a P.A.R.T.Y. program conducted in Australia^12^. A 10-year study conducted in Canada found that the effectiveness of the P.A.R.T.Y. program in reducing the incidence of trauma was significantly greater among female participants than male participants^14^. Possible explanations include lower interest or perceived relevance of the content among male students, as well as cultural embedded norms that encourage females to prioritize health and caregiving, while promoting traditional constructs of masculinity among males^15^. More than 63% of all injury-related deaths occurred among males, highlighting the urgent need for targeted injury prevention strategies to address gender disparities in awareness and motivation^1^.

The study findings may be influenced by a degree of selection bias, attributable to the underrepresentation of male participants and the relatively lower follow-up participation rate among Black students. Additionally, the questionnaire data may be subject to response bias, as participants’ answers could be affected by misinterpretation or social desirability. Nevertheless, the use of multiple measurement points likely mitigated these effects by maintaining relative consistency in any potential biases across assessments. Due to a substantial number of students not yet reaching the three-month follow-up window, data from the post-post survey were not used as the primary basis for conclusions. Although some studies have reported a significant attenuation of health education effects by four months, the available data in the current study do not demonstrate this trend^12^.

The YSL program demonstrated effectiveness in enhancing high school students’ injury awareness and response skills, with particularly notable improvements in bleeding injury management. Nevertheless, the observed variability in outcomes across demographic groups underscores persistent challenges in achieving equitable access and engagement.

## Data Availability

All data produced in the present study are available upon reasonable request to the authors

Data may be obtained from a third party and are not publicly available. The data that support the findings of this study are available from the corresponding author on reasonable request.

## Patient consent for publication

Not applicable.

## Ethics approval

The Quality Improvement and Risk Management Office of Sunnybrook Health Sciences Centre, Toronto, Canada waived ethical approval for this work, as the project was determined to be a Quality Improvement initiative and not human subject research. The project was registered internally under Quality Improvement Project Registration Number 1377951269.

## Acknowledgments

The authors acknowledge the assistance and support from staff of Sunnybrook Health Centre and department of Centre for Injury Prevention.

## Contributors

QM was involved in data analysis, interpretation of results, and drafting the manuscript. BT and SF conceptualized and oversaw the project and contributed to writing and revising the manuscript. BT serves as the guarantor for the manuscript.

## Reference

1. Global Health Estimates: Life expectancy and leading causes of death and disability. Accessed July 16, 2025. https://www.who.int/data/gho/data/themes/mortality-and-global-health-estimates

2. Government of Canada SC. Leading causes of death, total population, by age group. April 13, 2021. Accessed July 17, 2025. https://www150.statcan.gc.ca/t1/tbl1/en/tv.action?pid=1310039401

3. Injury costs across the lifespan – Parachute. Accessed August 29, 2025. https://parachute.ca/en/professional-resource/cost-of-injury-in-canada/injury-costs-across-the-lifespan/

4. Steinberg L. Risk Taking in Adolescence: New Perspectives From Brain and Behavioral Science. Curr Dir Psychol Sci. 2007;16(2):55–59. doi:10.1111/j.1467-8721.2007.00475.x

5. Steinberg L. Risk taking in adolescence: what changes, and why? Ann N Y Acad Sci. 2004;1021:51–58. doi:10.1196/annals.1308.005

6. Davison C. Injury Among Young Canadians: A National Study of Contextual Determinants.; 2014.

7. Pickett W, Molcho M, Simpson K, et al. Cross national study of injury and social determinants in adolescents. Inj Prev J Int Soc Child Adolesc Inj Prev. 2005;11(4):213–218. doi:10.1136/ip.2004.007021

8. Simpson K, Janssen I, Craig WM, Pickett W. Multilevel analysis of associations between socioeconomic status and injury among Canadian adolescents. J Epidemiol Community Health. 2005;59(12):1072–1077. doi:10.1136/jech.2005.036723

9. Michie S, van Stralen MM, West R. The behaviour change wheel: a new method for characterising and designing behaviour change interventions. Implement Sci IS. 2011;6:42. doi:10.1186/1748-5908-6-42

10. Schwarzer R. Modeling Health Behavior Change: How to Predict and Modify the Adoption and Maintenance of Health Behaviors. Appl Psychol. 2008;57(1):1–29. doi:10.1111/j.1464-0597.2007.00325.x

11. Ho KM, Litton E, Geelhoed E, et al. Effect of an injury awareness education program on risk-taking behaviors and injuries in juvenile justice offenders: a retrospective cohort study. PloS One. 2012;7(2):e31776. doi:10.1371/journal.pone.0031776

12. Cameron CM, Eley R, Judge C, O’Neill R, Handy M. Did attending P.A.R.T.Y. change youth perceptions? Results from 148 Queensland schools participating in the Prevent Alcohol and Risk-Related Trauma in Youth Program, 2018-2019. Inj Prev J Int Soc Child Adolesc Inj Prev. 2022;28(3):218–224. doi:10.1136/injuryprev-2021-044222

13. Brockamp T, Koenen P, Caspers M, et al. The influence of an injury prevention program on young road users: a German experience. Eur J Trauma Emerg Surg Off Publ Eur Trauma Soc. 2019;45(3):423–429. doi:10.1007/s00068-017-0872-9

14. Banfield JM, Gomez M, Kiss A, Redelmeier DA, Brenneman F. Effectiveness of the P.A.R.T.Y. (Prevent Alcohol and Risk-related Trauma in Youth) program in preventing traumatic injuries: a 10-year analysis. J Trauma. 2011;70(3):732–735. doi:10.1097/TA.0b013e31820783a3

15. Mikkonen J, Raphael D. Social Determinants of Health: The Canadian Facts. York University, School of Health Policy and Management; 2010.

